# Health services provided at the time of abortion in the US: a scoping review of the qualitative and quantitative evidence

**DOI:** 10.1101/2024.01.22.24301627

**Authors:** Katherine M Mahoney, Licia Bravo, Arden McAllister, Kacie Bogar, Sean Hennessey, Courtney A. Schreiber, Alice Abernathy

## Abstract

**Objectives:** While it is well documented that abortion access is associated with improved health, pregnancy-related, and socioeconomic outcomes, the association between abortion access and other reproductive health outcomes is less well described. Abortion-providing clinics also offer preventative reproductive health services. We conducted a scoping review to ascertain the extent to which preventive reproductive healthcare services (contraception, sexually transmitted infection testing and treatment, cervical cancer screening) are affected by abortion access in the United States.

**Methods:** Researchers screened articles and extracted data from PubMed, Embase, Scopus and CINAHL. We excluded articles that did not link abortion to contraception, sexually transmitted infection testing and treatment and cervical cancer screening; or took place outside the US.

**Results:** 5,359 papers were screened, 74 were included for full text review. Sixty-five were about contraception, seven on STIs, one on cervical cancer screening, and one on other services. The association between policies that restrict or protect abortion access and preventative health services has not been studied on a national scale. Drivers of variation were: insurance and billing policies; regulatory requirements of abortion-providing facilities, lack of staff training in clinics that did not specialize in abortion care; and limited follow up after abortion.

**Conclusions:** Abortion--providing clinics are a highly utilized access point for reproductive health services. More research is needed to determine the public health impact of constrained abortion access on contraceptive use, STI rates and cervical cancer in regions where many abortion-providing clinics have closed.

**Implications:** Attention should be paid to changing trends in contraceptive use, STI rates and cervical cancer as abortion-providing clinics close, this may reduce access to reproductive health services broadly.

## 1. Introduction

While state-level incursions on abortion provision created barriers to abortion care prior to the Dobbs v. Jackson Women’s Health Organization (Dobbs) decision in June 2022, the Dobbs decision precipitously reduced the number of abortion-providing clinics in the United States (US) (1, 2). State legislation restricting abortion, often resulting in abortion clinic closure, has outpaced legislation expanding abortion access (3).

Constrained abortion access affects public health through changes in birth rate, maternal morbidity and mortality, and provider training (4). Abortion clinics are a site for comprehensive reproductive healthcare delivery, including contraception (5–7), sexually transmitted infection (STI) testing and treatment (8, 9), cervical cancer screening (7, 8), and other preventative health services. Neither the distribution of contraception, STI testing and treatment, cervical cancer screening at abortion-providing clinics, nor the impact of abortion-related policy changes on these services have been systematically examined.

Without a synthesis of existing literature, understanding the full impact of policies that restrict or expand abortion access is limited. Thus, we conducted a scoping review to: (1) outline the current status of providing comprehensive reproductive healthcare at the time of abortion; (2) describe how policy changes have exacerbated existing barriers or facilitated delivery of these services at the time of abortion and (3) identify knowledge gaps regarding how this rapidly evolving policy landscape has affected delivery of other reproductive health services by abortion-providing clinics.

## 2. Materials and Methods

### 2.1 Protocol and registration

We followed the PRISMA guidelines for scoping reviews. We registered the protocol with Open Science Framework.

### 2.2 Scope

We aimed to examine the extent to which comprehensive reproductive health services, defined as contraception, STI screening and treatment, cervical cancer screening, are offered at US abortion-providing clinics, and highlight knowledge gaps in the geographic distribution of reproductive healthcare at the time of abortion. The key questions addressed in this review are:

1. What is the current status of contraception provision, STI screening and treatment, cervical cancer screening at the time of abortion or in clinics that provide abortion?
2. What are the effects of abortion policy changes or clinic closures on reproductive health services (contraception, STI testing, cervical cancer screening) in abortion-providing clinics?
3. Identify knowledge gaps regarding how clinic closures and policy changes have affected delivery of such care.

Articles were eligible for review if they related contraception, cervical cancer screening, STI screening/treatment/rates to abortion access, demand or provision. Articles on abortion access alone or those with a non-US focus were excluded. We did not restrict by year of publication or study design.

### 2.3 Search and Screening Strategy

We developed our search strategy in consultation with a librarian at the University of Pennsylvania, recommendations from team members with experience conducting scoping reviews, and published guidelines(10). We searched PubMed, Embase, Scopus and CINAHL from inception to March 24, 2023: (Table 1).

**Table 1:** Search strategy for each database performed inception to March 24, 2023.

| Database | Search Strategy | Number of Results |
| --- | --- | --- |
| PubMed<br>February 28,2023 | ((((pap smear) OR (sexually transmitted infection)) OR (contraception))) AND (abortion, induced)) AND ((((((closure) OR (abortion ban)) OR (state policy)) OR (Dobbs)) OR (access)) OR (utilization)) | 4,306 |
| Embase<br>March 12, 2023 | ('pap smear'/exp OR 'pap smear' OR (pap AND ('smear'/exp OR smear)) OR 'sexually transmitted infection':ti,ab,kw OR contraception:ti,ab,kw) AND ('abortion ban':ti,ab,kw OR 'state policy':ti,ab,kw OR dobbs:ti,ab,kw) OR 'abortion, induced':ti,ab,kw | 444 |
| Scopus<br>March 20, 2023 | (abortion) AND ((access) OR (abortion AND ban) OR (abortion AND bans) OR (closure) OR (state AND policy) OR (abortion AND restrictions) OR (Dobbs)) AND ((contraception) OR (sexually AND transmitted AND infection) OR (pap AND smear)) AND ((utilization) OR (use) OR (availability) OR (provision)) | 236 |
| CINAHL<br>March 24, 2023 | TX (abortion) AND ((access) OR (abortion AND ban) OR (abortion AND bans) OR (closure) OR (state AND policy) OR (abortion AND restrictions) OR (Dobbs)) AND | 373 |
|  | ((contraception) OR (sexually AND transmitted AND infection) OR (pap AND smear)) AND ((utilization) OR (use) OR (availability) OR (provision)) |  |

References were uploaded into Covidence (11). Each title and abstract was reviewed by two of the five reviewers. We required consensus between two reviewers for full text review. Conflicts were resolved by discussion and consensus among all reviewers. One of five authors reviewed the full text and abstracted data. An additional author reviewed the extracted data before it was finalized.

### 2.4 Data Extraction

We extracted study ID, title, authors, inclusion criteria, region of US, methods, aims, study design, start date, end date, participants, population, total participants, and patient-, public health-, and policy-related outcomes. We organized data by reproductive health service delivered at the time of abortion (contraception, STI testing/treatment, cervical cancer screening). We identified policy changes that affected the above-listed outcomes. We did not conduct a critical appraisal or synthesis of the study findings, as this is not recommended for scoping reviews.

## 3. Results

A total of 5,359 papers were eligible for initial review. After excluding duplicates, non-US studies, or that did not link contraception, STIs, or cervical cancer screening to abortion access, demand or provision, 74 papers were included for full text review (Fig 1). There were 65 papers on contraception, seven on STIs, one on cervical cancer screening, and one on other healthcare services. Some papers crossed categories (Table 2). Publication years ranged from 1974 to 2023 and designs included randomized control trials, cohort and cross-sectional studies, qualitative, systematic review, and opinion pieces.

**Figure 1.**
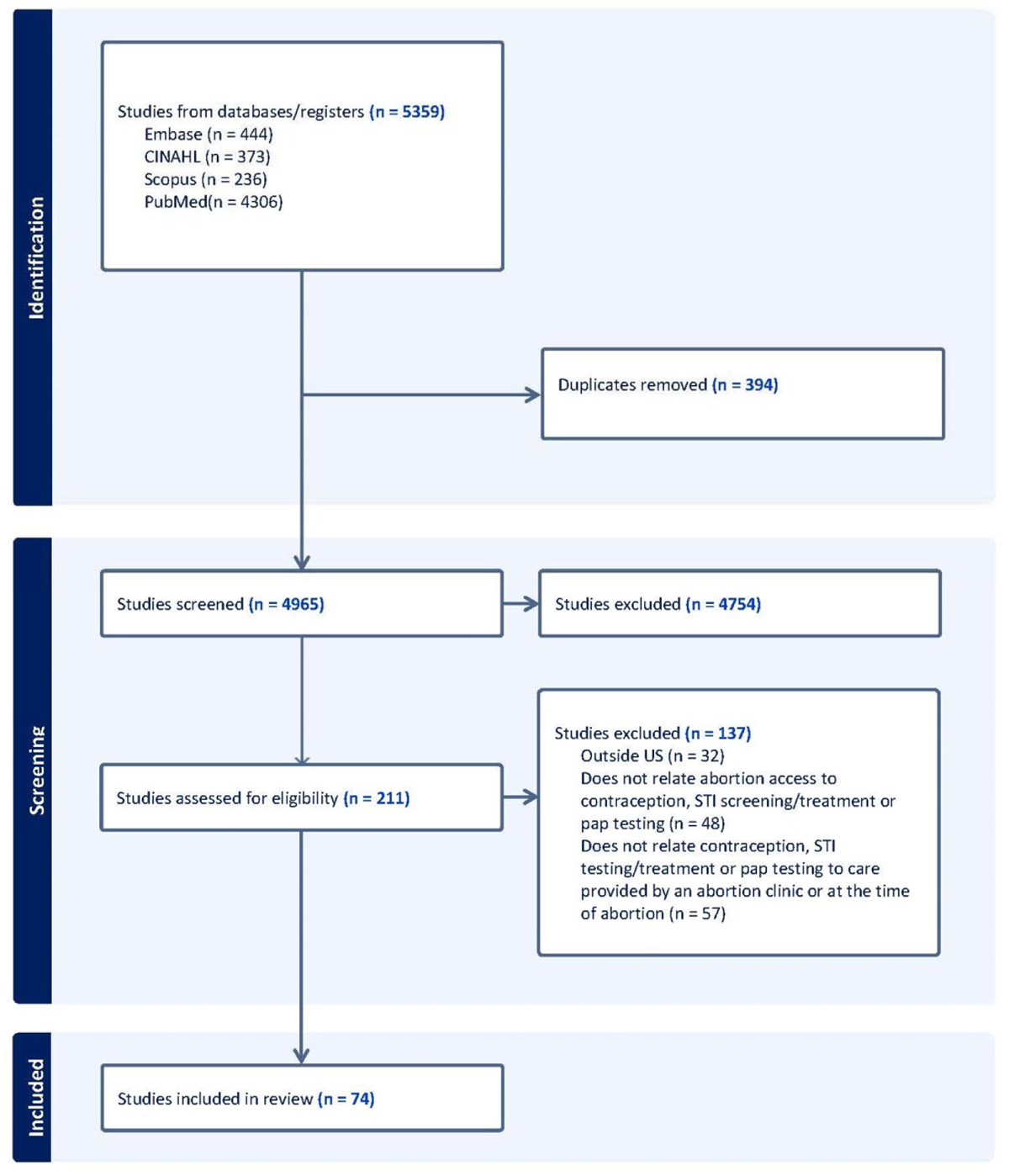
PRISMA diagram of studies screened and included in review.

**Table 2.**
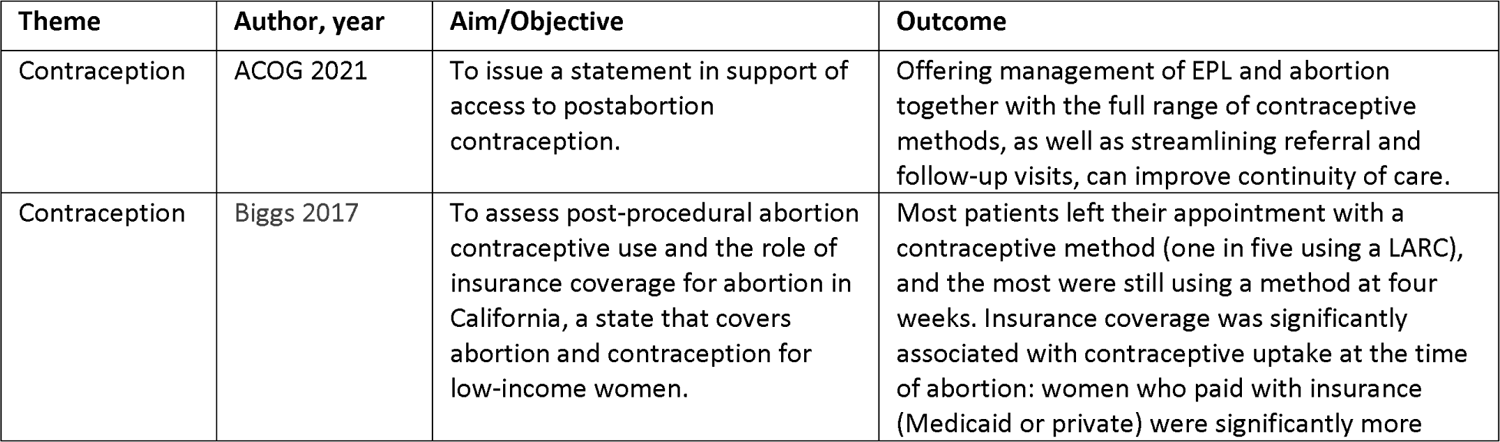
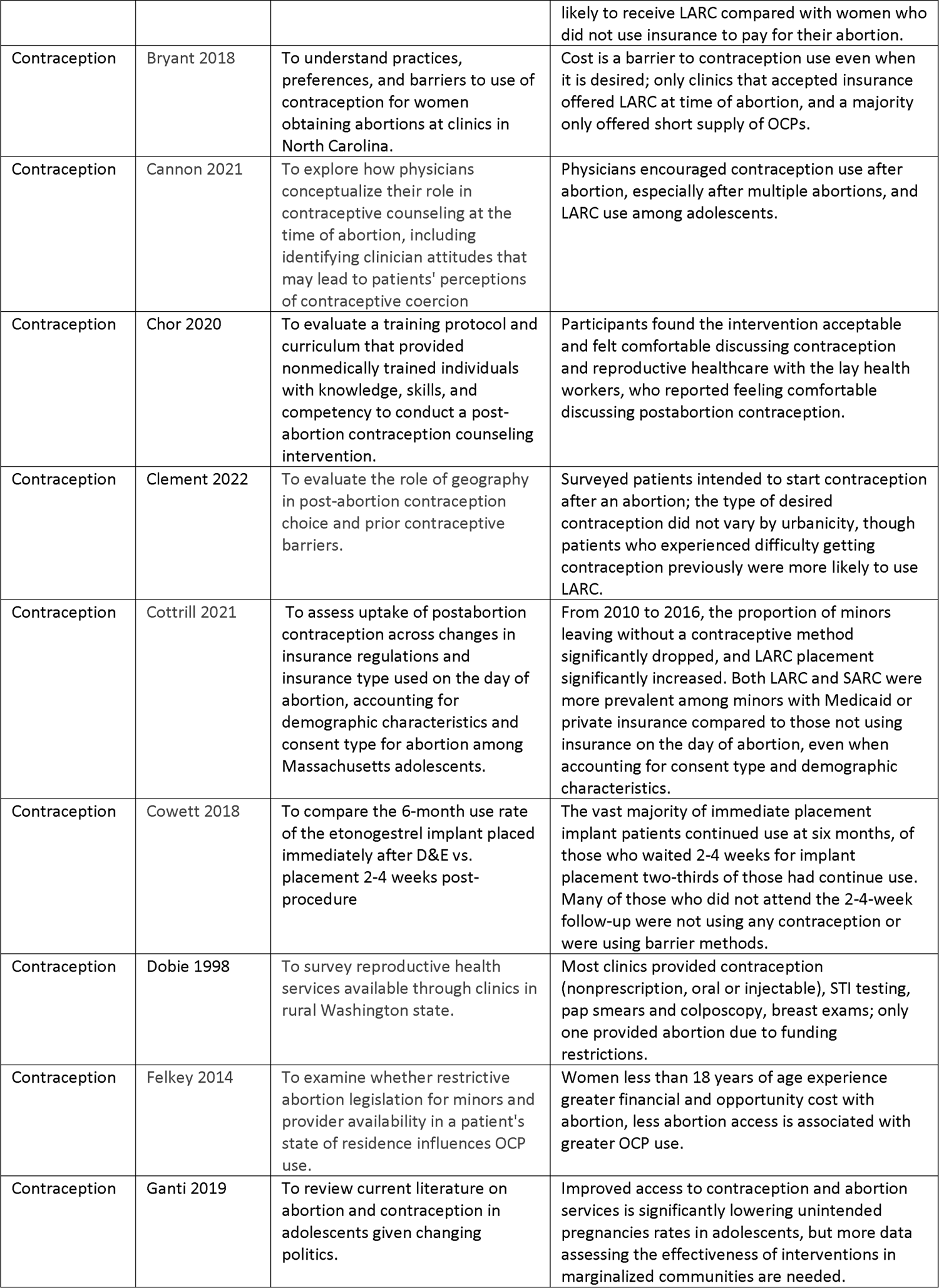
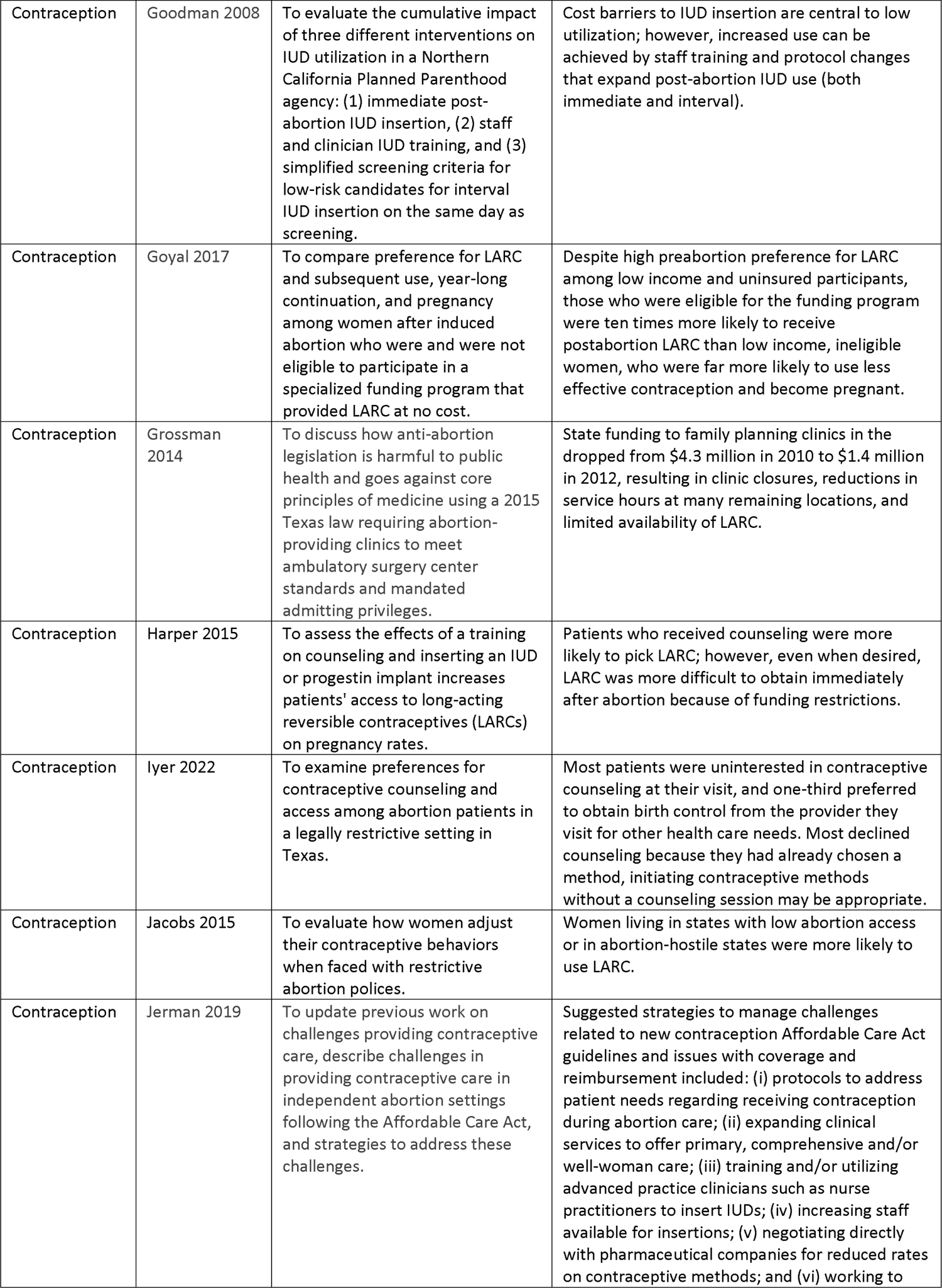
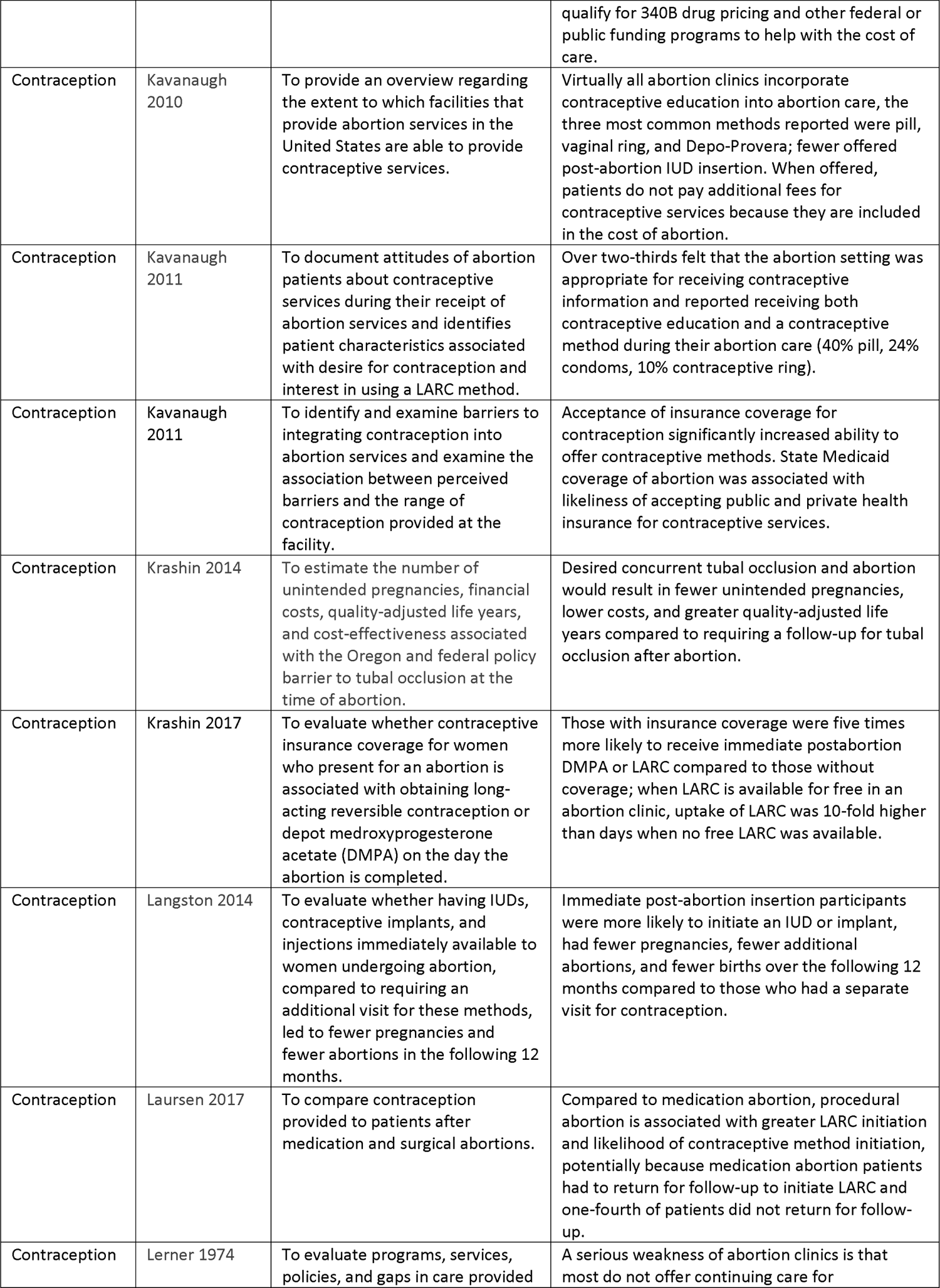
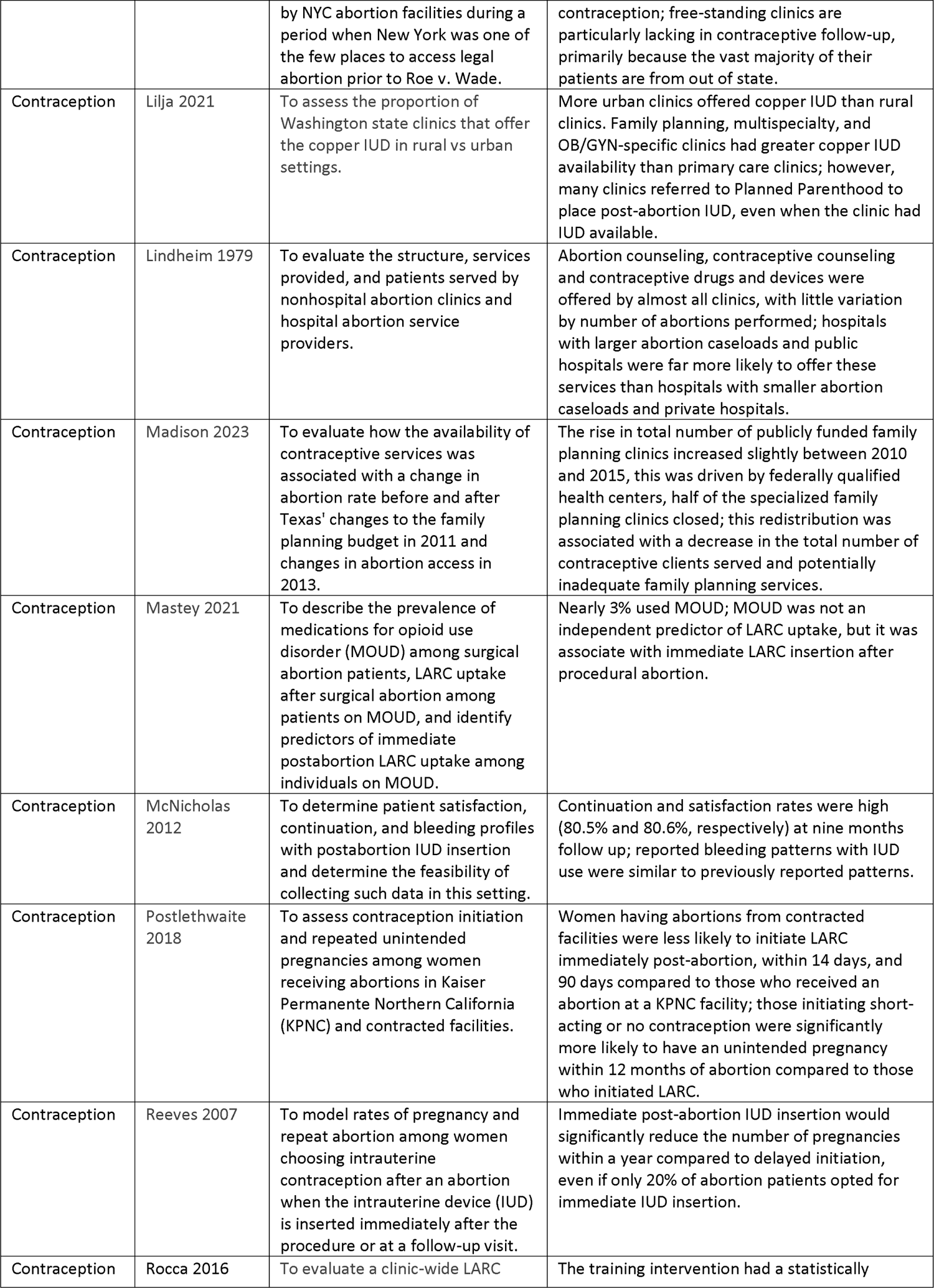
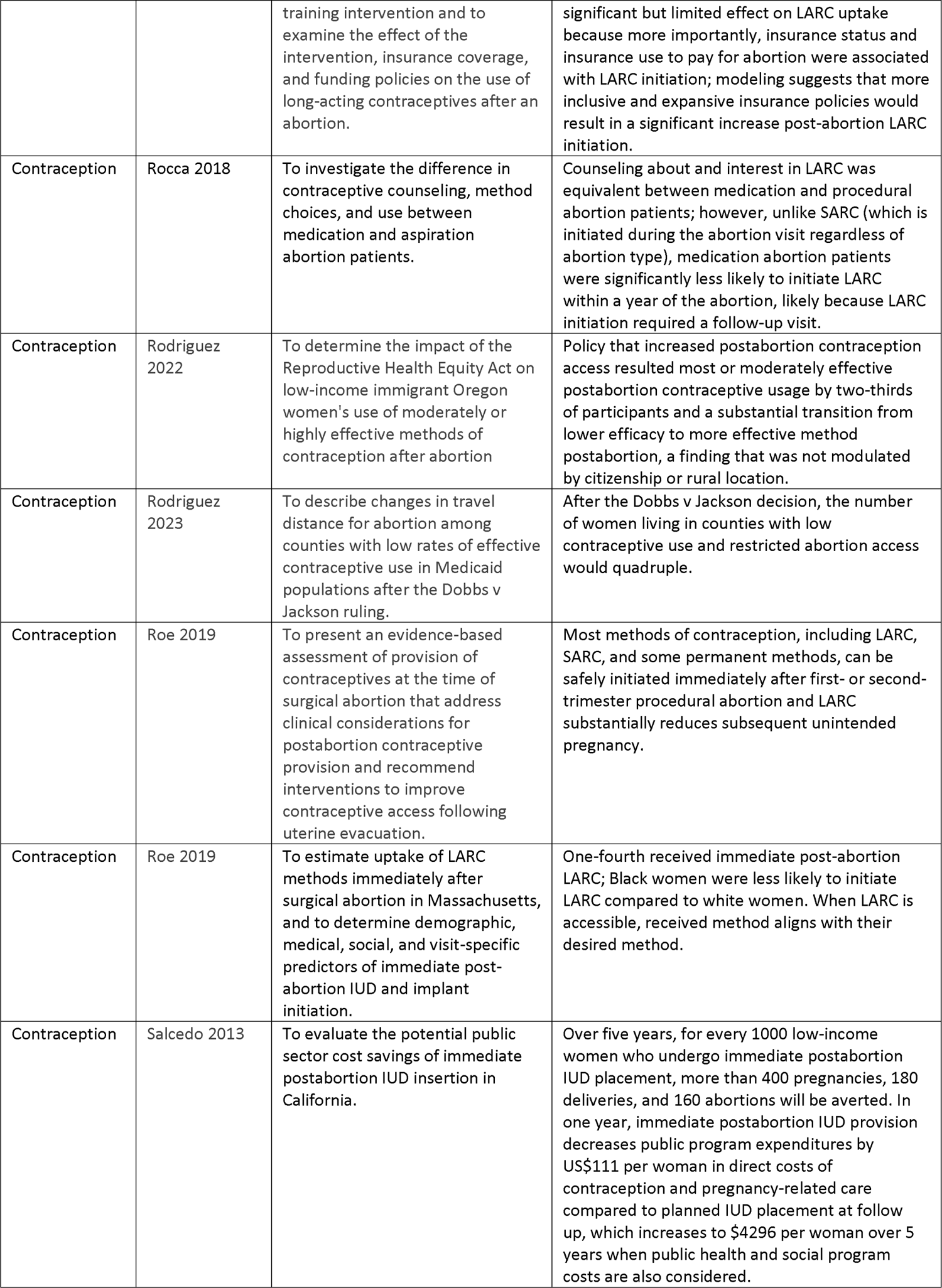
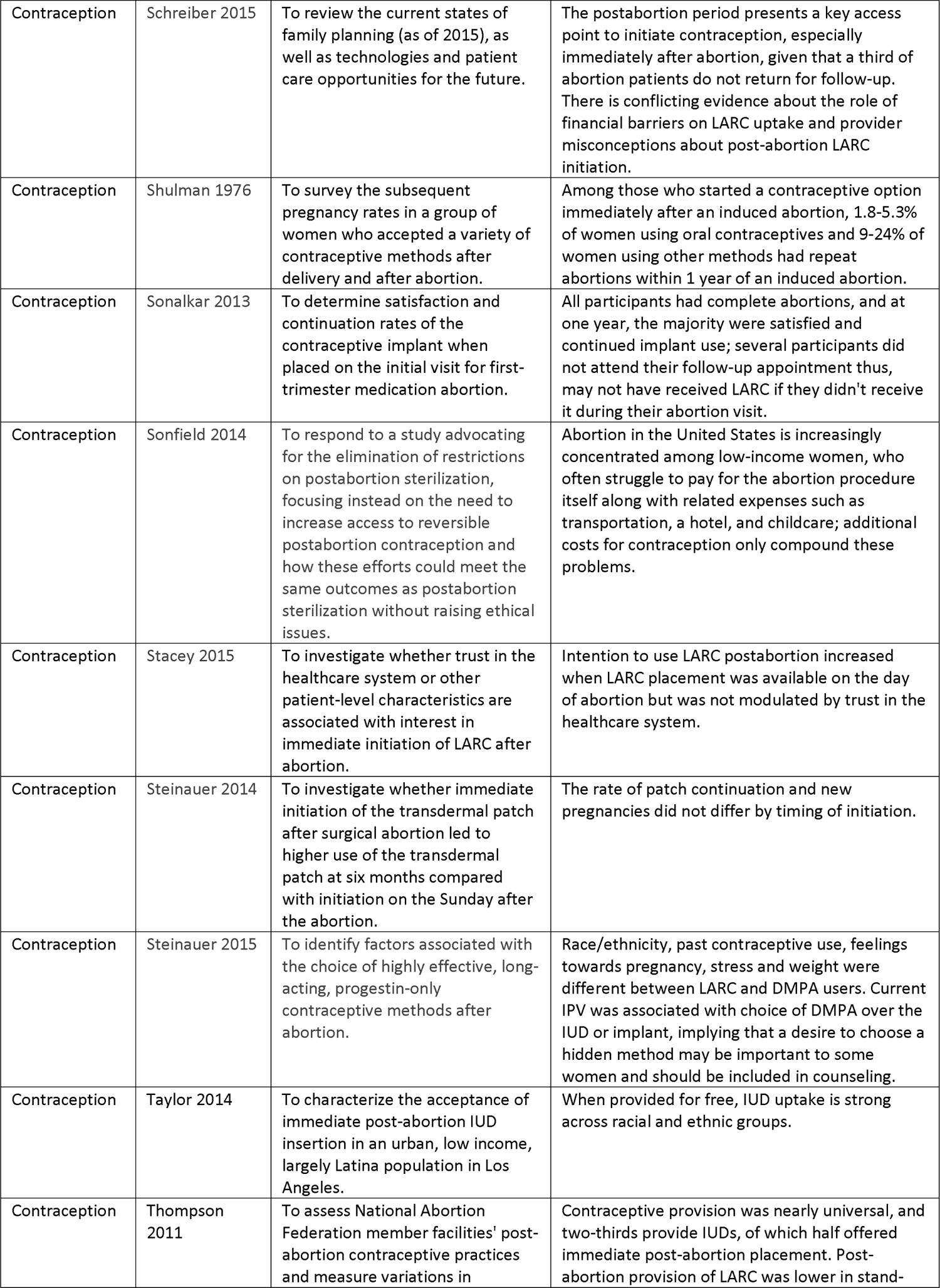
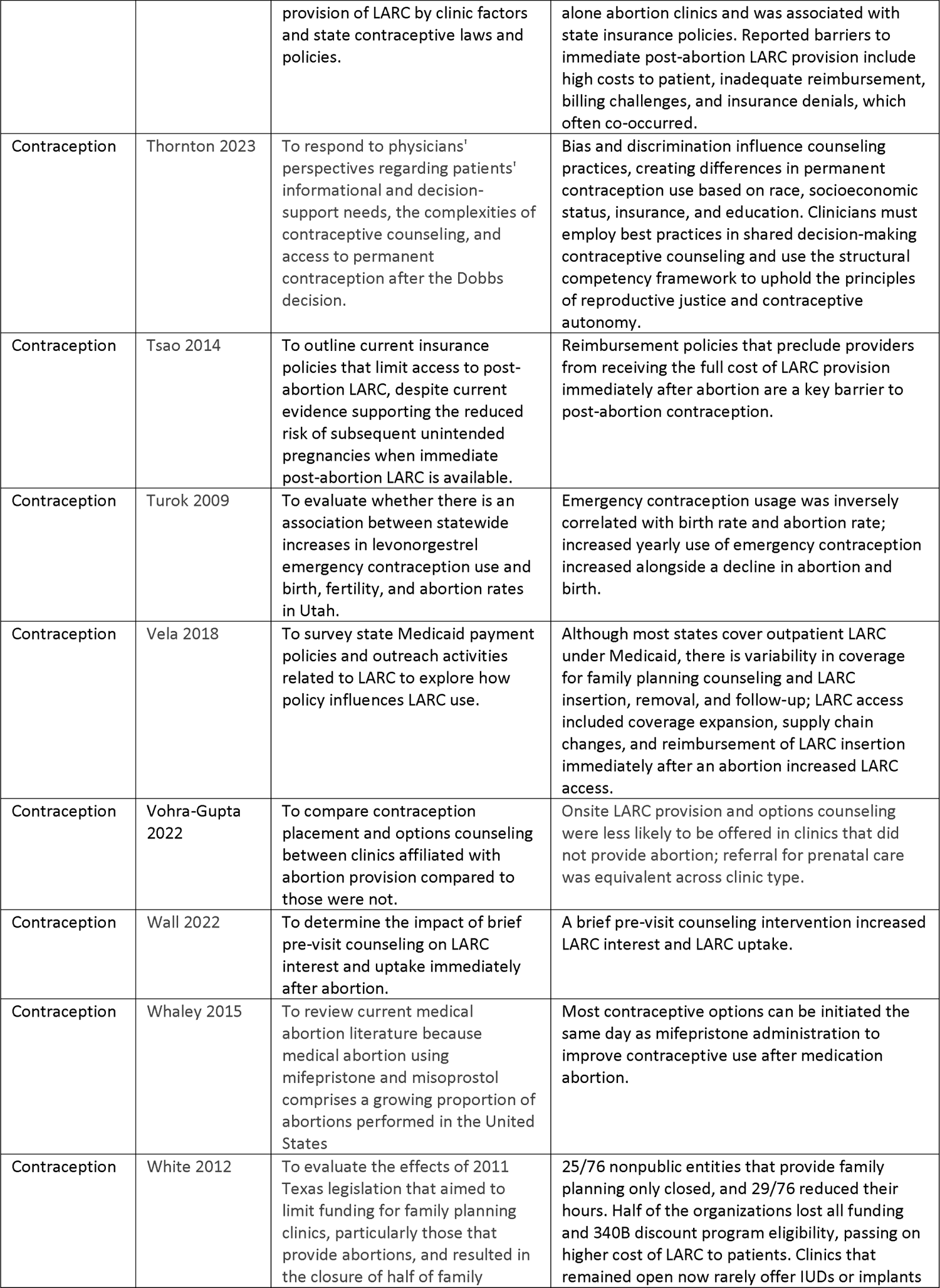
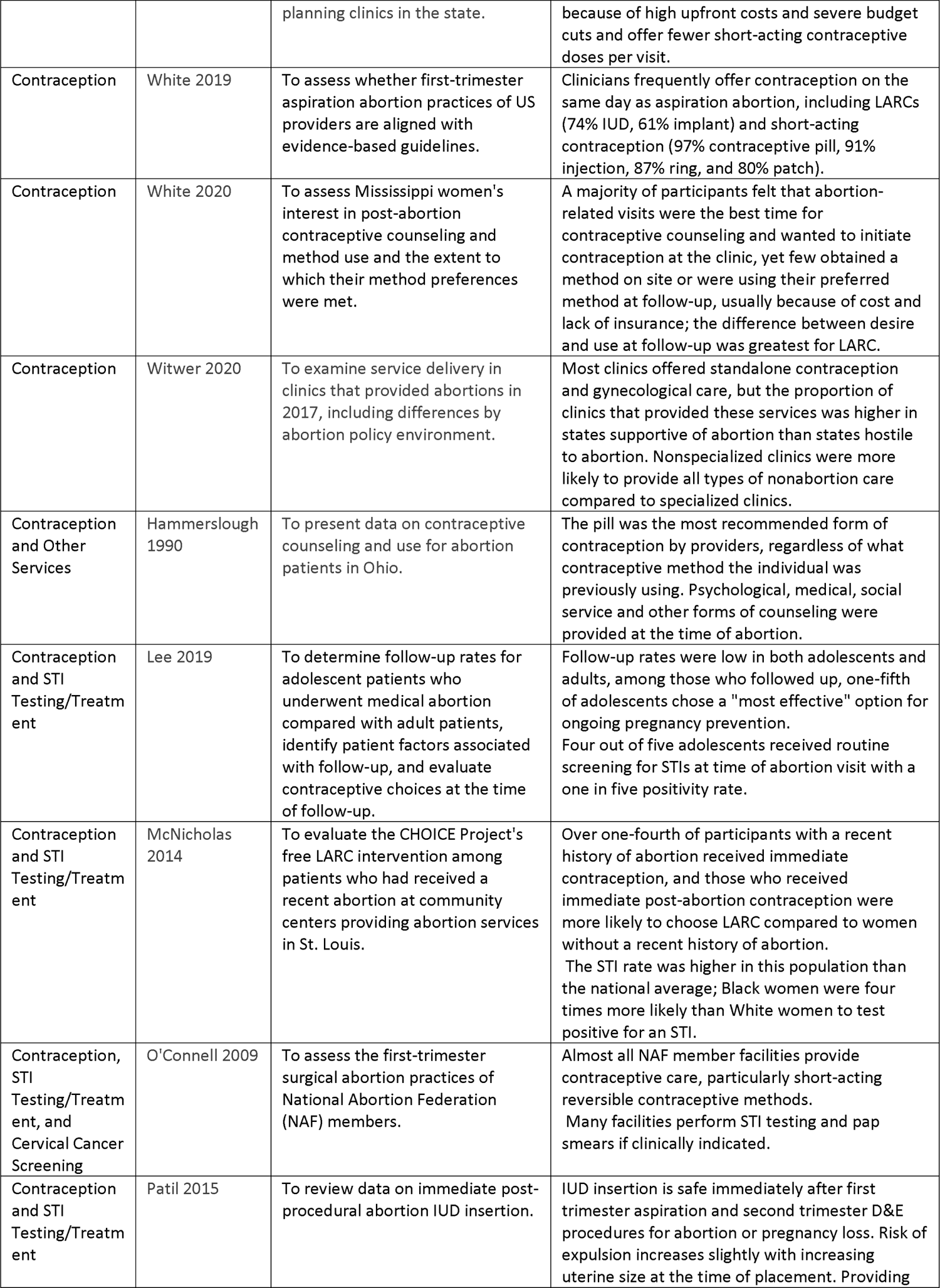
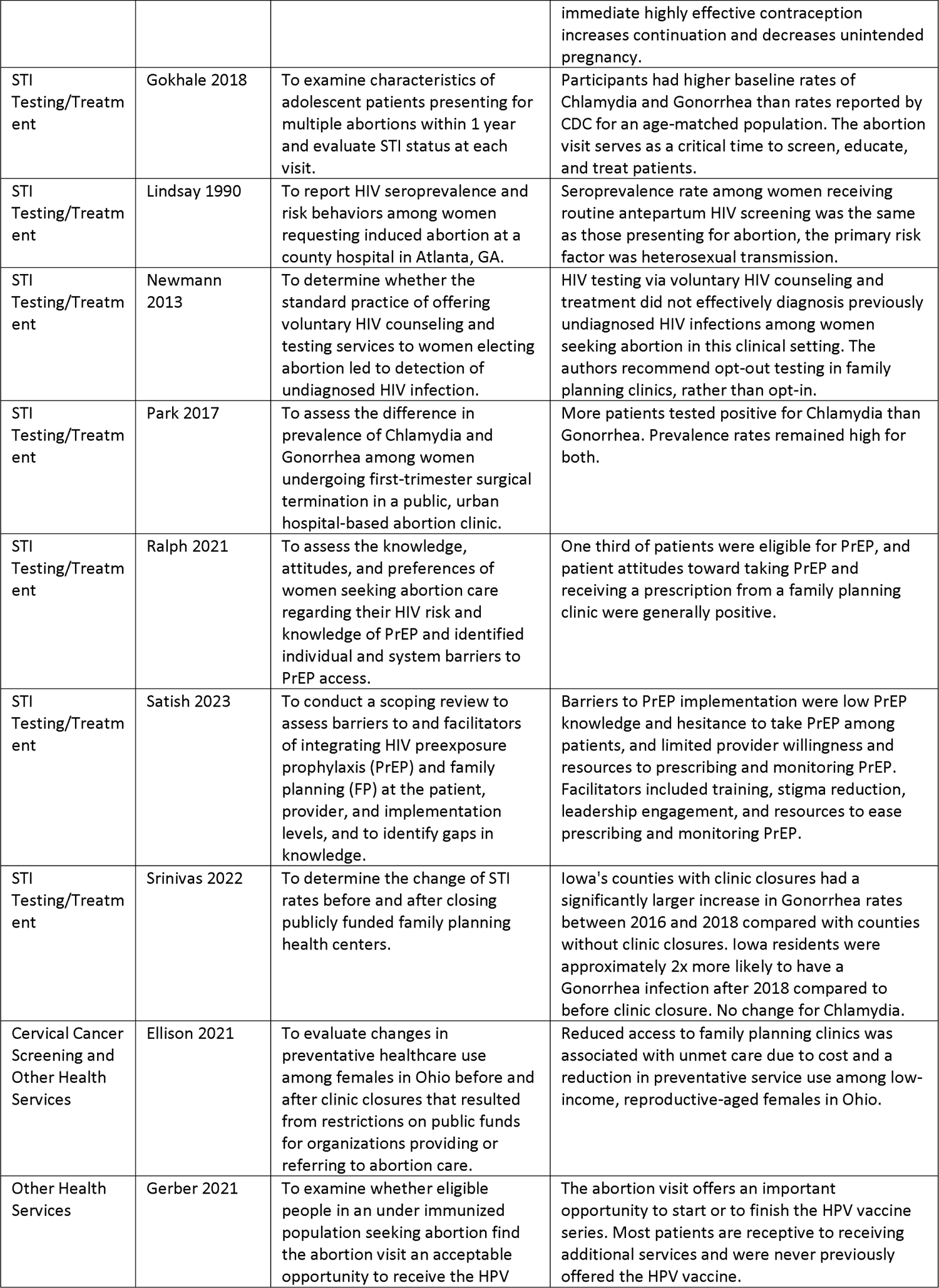
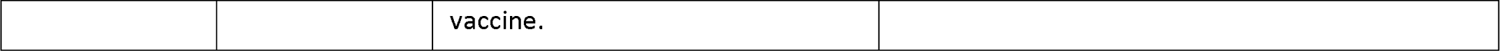
Papers that underwent full text review, organized by health service provided at abortion clinics and/or at the time of abortion.

### 3.1 Contraception

#### 3.1.1. Provision of contraception services at the time of abortion

Sixty-six studies examined contraceptive services. Most clinics offered contraceptive education, though methods were not available at all clinics (Kavanaugh 2010, O’Connell 2009, White 2019, Lerner 1974, Kavanaugh 2011, Roe 2019, Hammerslough 1990, Bryant 2018, Lindheim 1979, Lilja 2021). Clinics that primarily provided abortion care were more likely to offer IUD compared to primary care of general obstetric and gynecology clinics (Lilja 2021). Clinics that accepted insurance placed more long-acting reversible contraceptives (LARCs), though reimbursement for services associated with insertion, removal and follow up varied by insurance (Bryant 2018, Vela 2018). Policies that restricted reimbursement for abortion or care provided at the time of abortion were associated with reduced LARC provision (Bryant 2018, Lerner 1974, Thompson 2011, Cottrill 2021, Mastey 2021).

Immediate post-abortion LARC insertion was associated with reduced rapid repeat pregnancies, particularly in adolescents (Ganti et al, 2019). Models of immediate LARC insertion found over 70,000 unplanned pregnancies (Patil et al, 2015), and 20,000 repeat abortions could be prevented annually in the US if 20% of patients opted for immediate post-abortion LARC (Reeves et al 2007).

There was low follow up for contraception after abortion (Lerner 1974, Rocca 2018, Iyer 2022, Laursen 2017, McNicholas 2012), resulting in lower LARC uptake for patients who had medical compared to surgical abortions where LARC was more often placed at the time of abortion (Rocca 2018).

#### 3.1.2 Patient desire for & satisfaction with post-abortion contraception

Five studies examined patients’ desire for post-abortion contraception (Kavanaugh 2011, White 2020, Stacey 2015, Bryant 2018, Clement 2022). Patients viewed abortion facilities as an appropriate place to receive contraception counseling, especially for those without a longitudinal care provider (White 2020, Kavanaugh 2011). Of patients who declined contraceptive counseling, most did so because they either preferred to discuss contraception closer to home, with their primary care doctor, or because they already chose a contraception method (Iyer 2022). Satisfaction with LARC at the time of abortion appeared to be driven by cost and placement at the time of abortion (McNicholas 2014, McNicholas 2012, Sonalkar 2013).

#### 3.1.3 Factors that affected patient’s selection of contraception

Numerous factors influence contraception choice at the time of abortion: interpersonal violence, receipt of LARC specific counseling, sedation for abortion, and insurance coverage (Steinauer 2015, Biggs 2017, White 2020, Sonfield 2014, Cottrill 2021, Krashin 2017, Rocca 2016, White 2012, Steinauer 2015, Roe 2019, Roe 2019, Harper 2015, Wall 2022, Rocca 2018, Goyal 2017, Iyer 2022, Cottrill 2021, Mastey 2021, Thompson 2011).

Insurance and cost barriers limit post-abortion contraception access (White 2012, Biggs 2017, White 2020, Harper 2015, Sonfield 2014, Cottrill 2021, Roe 2019, Krashin 2017, Rocca 2016, Rocca 2018, Thompson 2011, Goyal 2017, Kavanaugh 2010, Bryant 2018, Kavanaugh 2011, Jerman 2019, Witwer 2020, Postlethwaite 2018, Vela 2018, Tsao 2014, Langston 2014, McNicholas 2014, Rodrigues 2022, Schreiber 2015, Taylor 2014). Insurance coverage was associated with a 30-100% increase in LARC uptake (Biggs, Rocca, Krashin Cottrill2021). Patients reported the primary reasons for not using a preferred contraceptive method were lack of insurance coverage and cost (White 2020). Patients may choose not use insurance for abortion-related care for confidentiality or logistical reasons (Sonfield 2014, Rocca 2016, Rocca 2018), and may be limited to more affordable contraceptives or forgo contraceptives (White 2012). Some (Roe 2019, Krashin 2017, Rocca 2016, McNicholas 2014, Goyal 2017, Rodrigues 2022, Taylor 2014) but not all (Schreiber 2015) found that when cost barriers are removed, effective contraception uptake increases.

Limited acceptance of insurance is a central obstacle in the provision of postabortion contraception (Thompson 2011, Kavanaugh 2010, Bryant 2018, Kavanaugh 2011, Sonfield 2014, Harper 2015, White 2012, Jerman 2019, Witwer 2020, Postlethwaite 2018, Tsao 2014, Langston 2014, Vela 2018). The percentage of clinics that do not accept insurance varied by clinic type and location (Kavanaugh 2010, Bryant 2018, Kavanaugh 2011). Reasons clinics did not accept insurance included reimbursement issues (Jerman 2019, Witwer 2020, Postlethwaite 2018, Tsao 2014, Sonfield 2014, Thompson 2011, Langston 2014, Vela 2018), high upfront cost of LARC (Sonfield 2014, Thompson 2011, Harper 2015, White 2012), and limited experience with billing insurance (Sonfield 2014).

Methods recommended to reduce insurance-related barriers included specialized funding programs for post-abortion contraception (Goyal 2017), negotiating directly with pharmaceutical companies for reduced rates on contraceptive methods (Jerman 2019), qualifying for 340B drug pricing and other federal or public funding programs (Jerman 2019), and hiring financial counselors to address patient-level barriers (Jerman 2019).

#### 3.1.4 Impact of abortion policy on contraceptive access and uptake

Restrictions on providing abortion care and decreased funding for family planning clinics disproportionately harmed residents in areas with already limited contraceptive access (Grossman et al. 2014). White et al. (2012) Access to the most effective contraceptive methods was reduced when abortion-providing clinics could not purchase contraceptives through 340B programs (White et al. 2012). Although the total number of family planning clinics in the form of federally qualified health centers increased in Texas, there was a decrease in contraceptive clients served (Madison et al. 2023).

Rodriguez et al. (2023) reported that the number of Medicaid enrolled females living in counties with low contraceptive use and restricted abortion access was expected to increase 46% following the Dobbs ruling.

Thompson et al. (2011) found that states with contraceptive coverage mandates and states with Medicaid family planning expansion program had higher rates of LARC use. Jacobs et al. (2015) found state-level abortion accessibility and hostility did not significantly modulate contraceptive use compared to individual characteristics.

### 3.2 STI testing and treatment

Lindsay and colleagues (1990) found that most patients were amenable to HIV testing at the time of abortion, and that universal screening was associated with improved detection among those without reported risk factors for HIV acquisition. Newmann et al recommended opt out counseling and treatment for HIV at the time of abortion (2013). Many abortion seeking patients had indications for HIV Pre-Exposure Prophylaxis (PrEP), though most expressed preference for starting PrEP with their PCP (Ralph et al, 2021). Satish et al. found other barriers to PrEP prescription at the time of abortion (2023).

Patients seeking abortion had a high prevalence of STIs (Park et al., 2017; Gokhale et al., 2018), and Black women were four times more likely than White women to have multiple STIs (McNicholas, 2014). Among adolescents seeking abortion, 80% were screened, with 22% testing positive (Lee 2019). Srinivas et al found increased incidence of gonorrhea in counties with an abortion clinic closure relative to those without a closure (Srinivas et al 2022).

### 3.3 Cervical cancer screening and prevention

Ellison et al found that restricted public funding of organizations that provide or refer for abortion was associated with reduced likelihood of receiving a pap (though this association was not as robust without imputation of key variables) (2021). O’Connell et al reported more than half abortion clinics perform STI testing and around half perform pap tests (2009).

### 3.4 Other public health services

Abortion clinics provide pelvic exams, pregnancy tests, hematologic assessments (Lindheim 1979), transgender-specific healthcare, mammogram and breast exams (Witwer et al., 2020) HPV vaccination (Gerber et al., 2021), and other forms of psychological, medical, and social service counseling (Hammerslough and Irizarry-Mora 1990). Ellison and colleagues (2021) found that in Ohio, restriction of public funds for abortion-providing clinics led to a decrease in likelihood of receiving a mammogram or breast exam; the effect was magnified as distance increased.

Mastey and colleagues (2021) observed the prevalence of opioid use disorder was twice the national average among abortion seeking patients (2.9 vs 1.5%), and proposed abortion clinics may present opportunities for opioid use screening and referral to care and treatment if desired.

## 4. Discussion

Findings from this scoping review suggest contraception, STI testing, and treatment and cervical cancer screening are acceptable to patients seeking abortion. Such care is consistent with guidelines endorsed by the Society of Family Planning and American College of Obstetricians and Gynecologists (5, 12). We anticipated variability in provision of contraception, STI testing and treatment and cervical cancer screening across abortion providing clinics and hypothesized the primary driver of variation of service provision was reimbursement difficulties and onerous operations requirements mandated at the state level. We did not find any national studies of the relationship between abortion policy and provision of contraception, STI testing and treatment, and cervical cancer screening. We found the most common drivers of variation in reproductive health service provision were: national and state insurance and billing policies that resulted in reimbursement challenges for care provided at the time of abortion, or by an abortion facility; facility capacity to provide services at the time of abortion; lack of staff training in clinics that did not specialize in abortion care; and limited patient follow up after abortion.

Abortion-providing clinics are a point of healthcare access for many patients who already experience structural and systemic barriers healthcare. For this reason, the impact of abortion-restrictive policies on access to other reproductive healthcare is not equally distributed among pregnancy-capable individuals. Populations particularly vulnerable to anti-abortion policies include adolescents, persons with limited financial resources, those who are under or uninsured, and those living in rural areas (Lindheim 1979, Felkey 2014, Reeves 2007, Dobie 1998, Lilja 2021). For these groups, care provided at the time of abortion may be one of the few times routine reproductive healthcare services are obtained. At the systems level, abortion care is often extensively regulated, and delivered through a combination of independent, state and federally funded clinics, which creates a complex health delivery network that is vulnerable to policy shifts that hinder abortion care provision and indirectly affect the concomitant services provided. As abortion care is restricted and abortion providing clinics close, patients that would obtain care at a clinic providing abortion will either forgo services or obtain them elsewhere. Whether the current infrastructure for care delivery is sufficient to meet this shifting demand remains unknown.

There are several limitations to this study. We did not consider global differences in abortion care. We conducted this review less than one year following Dobbs. There will likely be forthcoming evidence describing the impact of constrained abortion access on reproductive healthcare.

## 5. Conclusion

This review provides important insights into additional needs that reproductive aged persons will face as abortion-providing clinics close and disparity in access to abortion increases. Surprisingly, the impact of policies that restrict or protect abortion access on the provision of contraception, sexually transmitted infection testing and treatment, and cervical cancer provision has not been studied on a national scale. More research is needed to examine the public health impact of constrained abortion access resulting from clinic closures after Dobbs. Attention should be paid to changing trends in contraceptive use, STI rates and cervical cancer in regions where many abortion-providing clinics have closed, thus reducing access to reproductive health services broadly.

## Data Availability

All data produced in the present work are contained in the manuscript.

## Notes

**Funding sources:** Dr. Abernathy is supported by a grant from the NIH (K12-HD001265)

### Competing Interest Statement

The authors have declared no competing interest.

### Clinical Protocols

https://osf.io/69bcr

### Funding Statement

Dr. Abernathy is supported by a grant from the NIH (K12-HD001265).

